# Low-Cost 3D-Printed Molds for PMMA Cranioplasty: Case Series and Workflow Analysis

**DOI:** 10.64898/2026.04.02.26349771

**Authors:** Tomas Gondra, Romina Gimbatti, Pablo Santangelo

## Abstract

**BACKGROUND:** Cranioplasty is an essential procedure to restore cranial integrity, protect neural structures, and improve cosmetic outcomes. However, commercially available implants are often costly, limiting their accessibility in public healthcare systems. Three-dimensional (3D) printing offers a low-cost alternative for producing patient-specific solutions.

**METHODS:** A retrospective case series of eight patients undergoing cranioplasty using customized polymethylmethacrylate (PMMA) implants fabricated with 3D-printed molds was conducted. Computed tomography (CT) scans were used for segmentation and digital modeling. Patient-specific molds were designed and printed preoperatively. Variables analyzed included design time, printing time, intraoperative workflow, and clinical outcomes.

**RESULTS:** Design time ranged from approximately 1 hour for small defects to 3 hours for larger defects. Printing time ranged from 2–3 hours for smaller defects and up to 8–10 hours for larger reconstructions. Satisfactory aesthetic outcomes were achieved in 7 of 8 patients (87.5%). No major implant-related complications were observed.

**CONCLUSION:** Low-cost 3D printing for PMMA cranioplasty is a feasible, accessible, and effective technique for cranial reconstruction, particularly in resource-limited settings.

## INTRODUCTION

Cranioplasty is a commonly performed neurosurgical procedure aimed at restoring cranial defects following trauma, tumor resection, or decompressive craniectomy. In addition to its protective role, cranioplasty has been associated with improved neurological recovery and cosmetic outcomes [1]. Despite the availability of various reconstructive materials, including titanium, polyetheretherketone (PEEK), and prefabricated polymethylmethacrylate (PMMA) implants, their high cost and limited accessibility remain significant barriers in public healthcare systems, particularly in low- and middle income countries. As a result, alternative strategies that balance affordability, customization, and surgical efficiency are needed. Recent advances in three-dimensional (3D) printing have enabled the development of patient-specific solutions at a substantially lower cost [2,3,4]. In this context, the use of 3D-printed molds for intraoperative PMMA implant fabrication represents a practical and accessible approach. The aim of this study is to present a case series of customized PMMA cranioplasties using low-cost 3D-printed molds and to evaluate their feasibility, workflow, and clinical outcomes.

## MATERIALS AND METHODS

### Study Design

A retrospective case series of consecutive patients undergoing cranioplasty using customized polymethylmethacrylate (PMMA) implants fabricated with 3D-printed molds was conducted.

### Study Setting and Patients

The study was performed at a tertiary public hospital. Eight consecutive patients with cranial defects of post-traumatic origin were included. Patients were selected based on the presence of a cranial defect requiring reconstruction and the feasibility of preoperative 3D planning. No patients were excluded once the technique was adopted.

### Imaging Acquisition and Processing

All patients underwent preoperative volumetric computed tomography (CT) scanning. DICOM data were imported into dedicated software for segmentation and 3D reconstruction of cranial anatomy. The cranial defect was digitally reconstructed, and a patient-specific mold was designed based on the expected anatomical contour.

### 3D Modeling and Printing

Digital models were processed to create negative molds suitable for intraoperative PMMA shaping. The molds were fabricated using low-cost fused deposition modeling (FDM) 3D printers. Printing parameters were adjusted according to defect size and complexity. The final printed molds were prepared and sterilized according to institutional protocols. The workflow for implant design, segmentation, and fabrication is illustrated in Figure 1.

**FIGURE 1:**
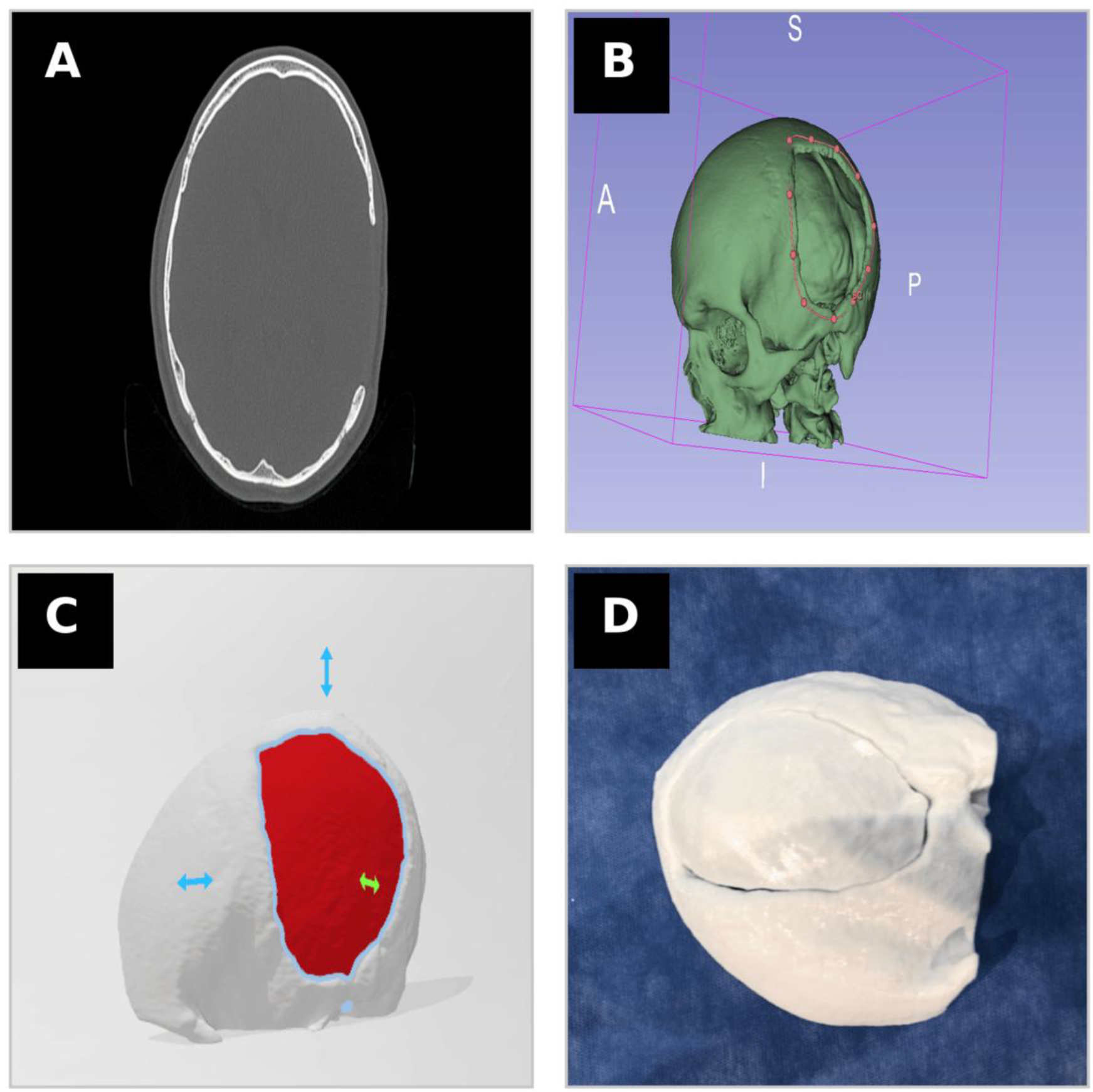
Workflow for customized cranioplasty using 3D-printed molds. (A) Axial CT scan demonstrating the cranial defect. (B) Three-dimensional reconstruction with segmentation and definition of defect margins. (C) Digital modeling of the patient-specific implant and mold. (D) Final 3D-printed mold prior to intraoperative use.

### Surgical Technique

Standard cranioplasty procedures were performed. PMMA was prepared intraoperatively and shaped using the customized 3D-printed molds to achieve accurate anatomical reconstruction. The implant was then positioned and fixed using standard fixation techniques.

### Variables and Outcomes

The following variables were analyzed: preoperative design time, printing time, intraoperative handling and need for adjustments, aesthetic outcome (subjective surgeon assessment), and perioperative complications. Data were analyzed descriptively. Continuous variables are presented as ranges, and categorical variables as proportions. No statistical comparisons were performed due to the small sample size.

## RESULTS

A total of eight patients underwent cranioplasty using customized PMMA implants fabricated with 3D-printed molds.

### Workflow Metrics

The preoperative digital design time varied according to defect size, averaging approximately 1 hour for smaller defects and up to 3 hours for larger or more complex defects. Similarly, 3D printing time ranged from 2-3 hours for smaller defects to 8-10 hours for larger reconstructions.

### Intraoperative Findings

The use of patient-specific molds allowed accurate shaping of the PMMA implants with minimal need for intraoperative adjustments. Implant fitting was considered satisfactory in all cases. The technique facilitated a streamlined intraoperative workflow by reducing manual modeling and improving reproducibility. The intraoperative fabrication and placement of the implant are illustrated in Figure *2*.

**FIGURE 2:**
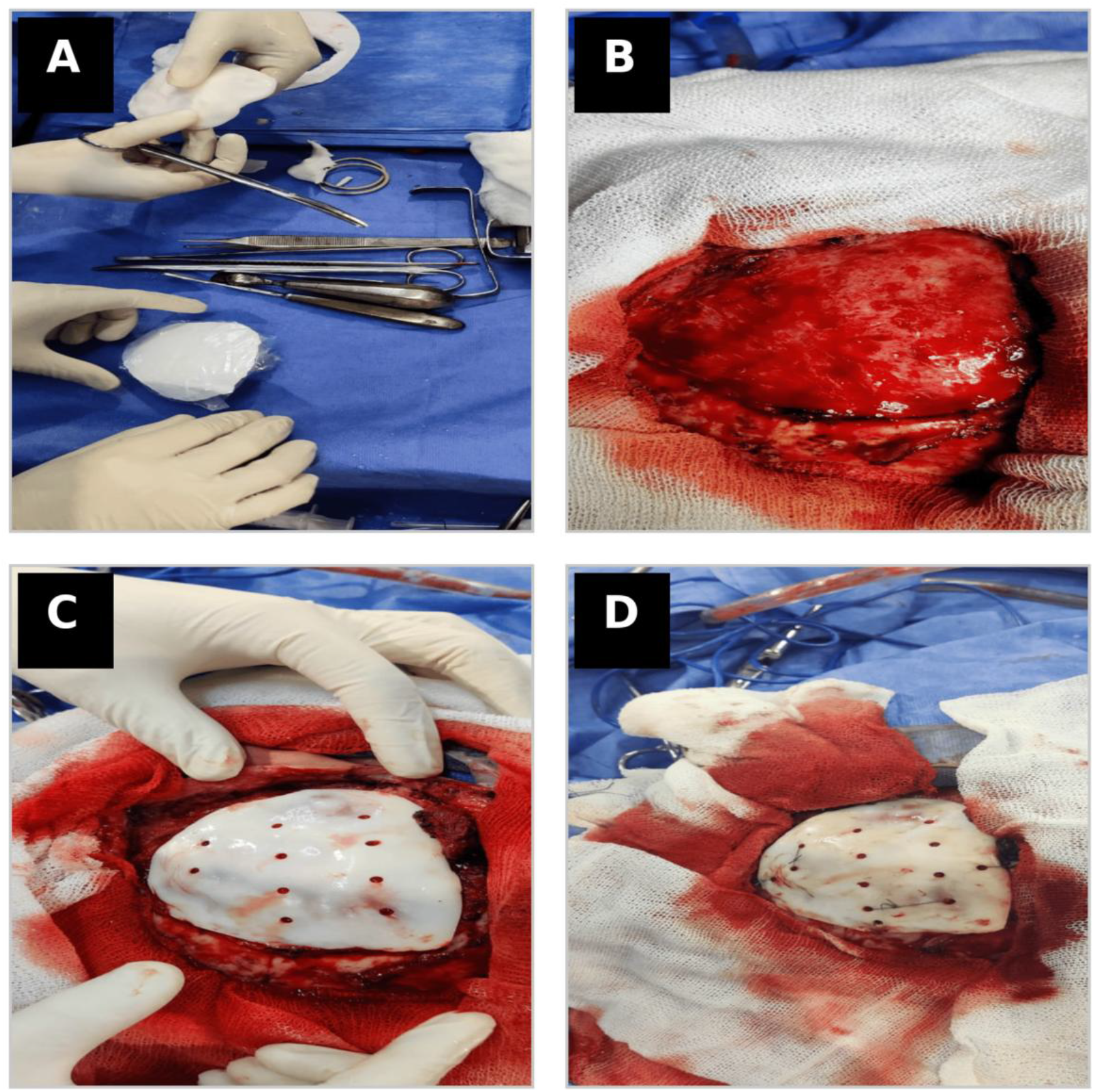
Intraoperative fabrication and placement of patient-specific PMMA implant. (A) Intraoperative preparation and handling of the PMMA implant (B) Intraoperative view of the cranial bone defect after exposure. (C) Implant positioned within the cranial defect. (D) Final fixation of the implant.

### Clinical Outcomes

Satisfactory aesthetic outcomes were achieved in 7 out of 8 patients (87.5%), based on the operating surgeon’s subjective assessment. Postoperative CT outcomes are illustrated in Figure *3*. Clinical photographs were not included due to the difficulty of ensuring complete patient anonymity while adequately demonstrating surgical outcomes. Therefore, imaging findings were prioritized for objective evaluation.

**Figure 3.**
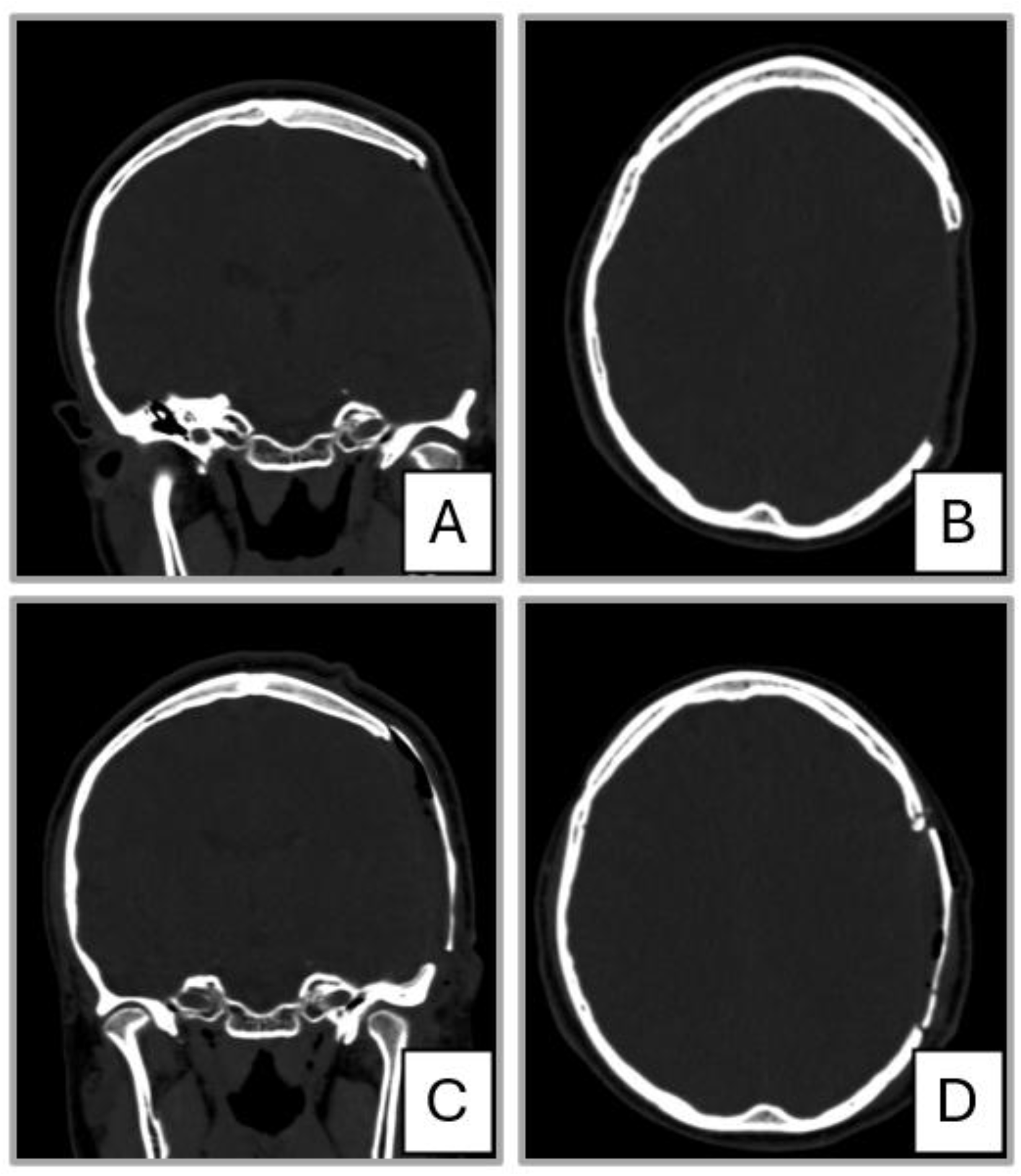
Preoperative and postoperative CT evaluation of cranial reconstruction. (A) Preoperative coronal CT demonstrating the cranial defect. (B) Preoperative axial CT showing the extent of the defect. (C) Postoperative coronal CT demonstrating restoration of cranial contour. (D) Postoperative axial CT showing adequate implant positioning.

No major implant-related complications were observed during the perioperative period. No postoperative infections or implant removals were recorded.

A summary of workflow metrics and clinical outcomes is presented in Table *1*.

**TABLE 1:**
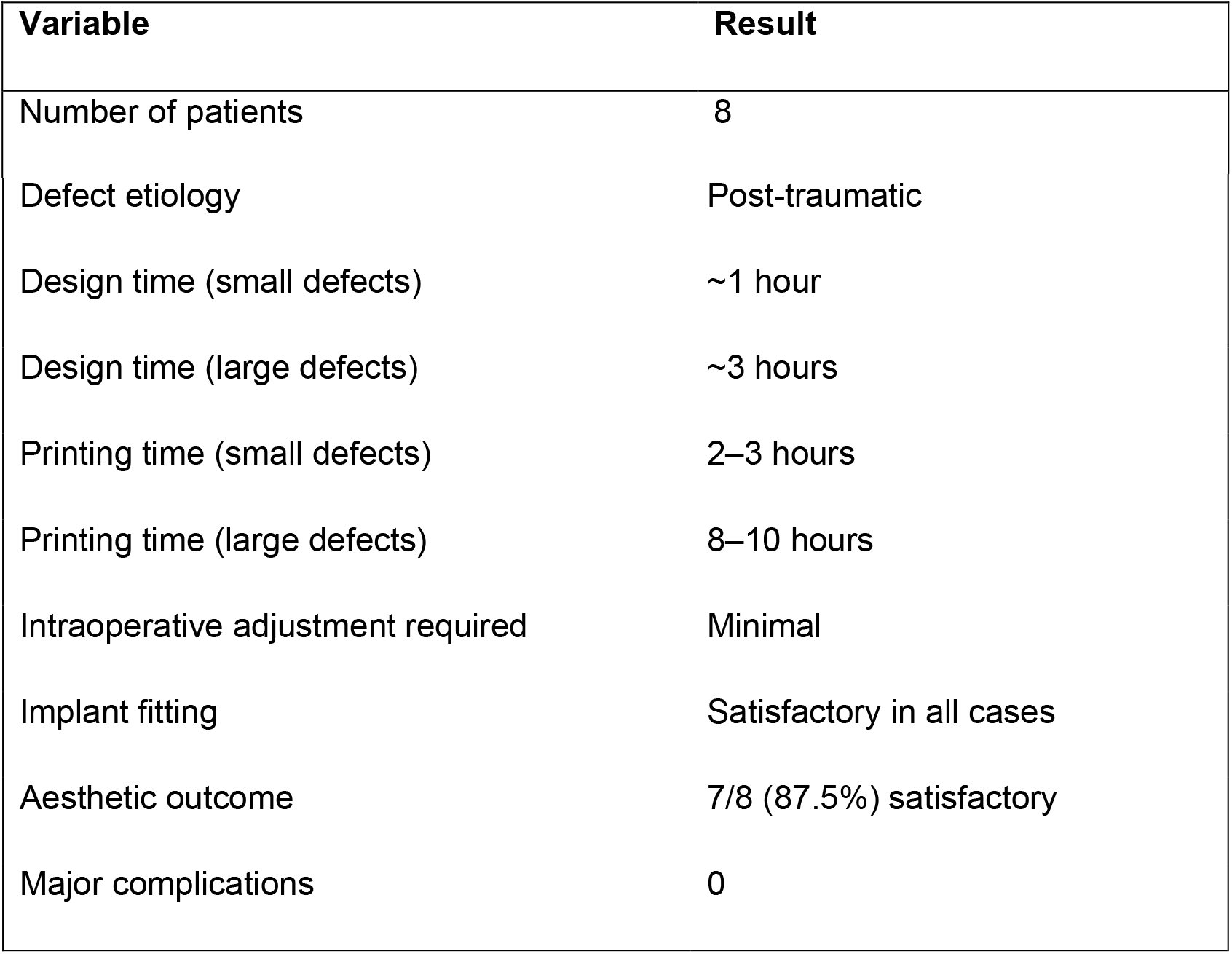
Workflow metrics and clinical outcomes of patients undergoing cranioplasty using 3D-printed molds. Values are presented as ranges or proportions. Aesthetic outcomes were assessed subjectively by the operating surgeon. No statistical analysis was performed due to the small sample size. Defect size classification was based on intraoperative assessment.

| Variable | Result |
| --- | --- |
| Number of patients | 8 |
| Defect etiology | Post-traumatic |
| Design time (small defects) | ~1 hour |
| Design time (large defects) | ~3 hours |
| Printing time (small defects) | 2–3 hours |
| Printing time (large defects) | 8–10 hours |
| Intraoperative adjustment required | Minimal |
| Implant fitting | Satisfactory in all cases |
| Aesthetic outcome | 7/8 (87.5%) satisfactory |
| Major complications | 0 |

## DISCUSSION

The use of low-cost 3D printing in cranioplasty has emerged as a valuable alternative to commercially available implants, particularly in settings with limited resources. Our experience supports the feasibility and practicality of this approach, demonstrating that patient-specific PMMA implants can be produced with satisfactory aesthetic and clinical outcomes. Previous studies have demonstrated the viability of low-cost 3D-printed solutions for cranial reconstruction [2,3]. Our findings are consistent with these reports, reinforcing the concept that technological accessibility has significantly expanded the possibilities for personalized neurosurgical care. One of the main advantages observed in our series is the improvement in implant accuracy and fit. Traditional freehand intraoperative modelling of PMMA can be time-consuming and highly dependent on surgeon experience. In contrast, the use of a predesigned mold allows for more predictable results, reduces intraoperative manipulation, and improves reproducibility. [4] From a practical standpoint, the trade-off between increased preoperative planning time and reduced intraoperative complexity appears favourable. In our series, design and printing times were reasonable and could be integrated into routine surgical workflows without delaying patient care. Another relevant aspect is cost-effectiveness. Previous studies comparing autologous bone flaps and synthetic implants have highlighted the economic and clinical implications of cranioplasty materials [3,5,6]. This approach is particularly relevant in low- and middle-income settings, where access to commercially available implants remains limited. In this context, low-cost 3D printing represents a particularly attractive alternative, especially in public healthcare systems where access to prefabricated implants is limited. [4,7] Despite these advantages, this study has several limitations. First, the sample size is small, which limits the generalizability of the findings. Second, the absence of a control group prevents direct comparison with other reconstruction techniques. Third, aesthetic outcomes were assessed subjectively, without standardized objective measurements. Finally, the successful implementation of this technique depends on the availability of basic technological resources and a certain degree of technical expertise. However, as 3D printing becomes increasingly accessible, its integration into routine neurosurgical practice is likely to expand.

## CONCLUSIONS

Customized PMMA cranioplasty using low-cost 3D-printed molds is a safe, feasible, and cost-effective technique for cranial reconstruction. This approach allows accurate implant fabrication, improves intraoperative workflow, and provides satisfactory aesthetic outcomes. Its implementation may be particularly valuable in resource-limited settings, where access to commercially manufactured implants is restricted. Further studies with larger cohorts and comparative designs are required to validate these findings. This technique represents a practical and scalable alternative for patient specific cranial reconstruction in public healthcare systems.

## Data Availability

All data produced in the present study are available upon reasonable request to the authors

## ADDITIONAL INFORMATION

All authors have reviewed the final version to be published and agreed to be accountable for all aspects of the work.

Human subjects: Informed consent for treatment and open access publication was obtained or waived by all participants in this study. CODEI Hospital Santojanni issued approval Not applicable. Ethics Statement This study was conducted in accordance with institutional ethical standards and the principles of the Declaration of Helsinki. The study protocol was reviewed and approved by the ethics committee of Hospital Santojanni.

Due to institutional administrative limitations, a formal approval identification number was not issued. Written informed consent was obtained from all patients for both treatment and publication purposes. No identifying patient information is included in this manuscript.

Conflicts of interest: In compliance with the ICMJE uniform disclosure form, all authors declare the following: Payment/services info: All authors have declared that no financial support was received from any organization for the submitted work. Financial relationships: All authors have declared that they have no financial relationships at present or within the previous three years with any organizations that might have an interest in the submitted work. Other relationships: All authors have declared that there are no other relationships or activities that could appear to have influenced the submitted work.

## Acknowledgements

The authors would like to acknowledge the technical support provided in the 3D modeling and printing process. The authors also acknowledge the institutional support that made this work possible. Data supporting the findings of this study are available from the corresponding author upon reasonable request. All data are de-identified to protect patient confidentiality.

